# A systematic scoping review on the impact of the COVID-19 quarantine on the psychological wellbeing of medical students

**DOI:** 10.1101/2021.11.28.21266956

**Authors:** Divya I. Vythilingam, William U. Atiomo

## Abstract

**Objectives:** The goal of this study was to identify the nature and extent of the available published research on the impact of social isolation, on the psychological wellbeing of medical students, who had to quarantine due to the COVID-19 pandemic.

**Design:** Scoping review.

**Search strategy:** The PRISMA-ScR (Preferred Reporting Items for Systematic reviews and Meta-Analyses extension for Scoping Reviews), guideline, was used to structure this study. A search strategy was carried out across six bibliographic databases. PubMed, Embase, ERIC, Scopus, Cochrane Database of Systematic Reviews and Web of Science. The following search terms were used, “medical student*” AND “impact” AND “quarantine” AND “COVID-19”. Searches were confined to articles published (excluding conference abstracts) between 1 January 2019-21 August 2021. A search of secondary references was conducted. Data from the selected studies were extracted, and the following variables recorded; first author and year of publication, country of study, study design, sample size, focus group, mode of analyzing impact of quarantine from COVID-19 on mental health and results of the studies.

**Results:** A total of 223 articles were identified across the six databases, from which 69 duplicates were excluded resulting in 154 full-text articles. Of these, 29 met the inclusion criteria. Following a review of the abstracts of these 29, ten full-text articles were identified all of which were cross sectional studies. Sample sizes ranged from 182 to 860 students and all studies used a variety of self-administered questionnaires to measure psychological wellbeing. Eight of the 10 articles showed that quarantine had a negative impact on the psychological well-being of medical students.

**Conclusion:** The evidence is small but growing. Quarantine because of the COVID-19 pandemic appears to have had a negative impact on the psychological wellbeing of medical students. There is a need for more studies to further evaluate this research question.

**Article summary:** Strengths and limitations of this study.

1. This was the first scoping review as far as we know of the literature in this area.
2. Many of the studies analysed in this review were cross-sectional in nature.
3. A variety of measurement tools were used to assess psychological wellbeing, preventing comparison of results and data synthesis.
4. Many of the studies included in the review did not include a control group of medical students, pre-COVID-19 quarantine.
5. A detailed critical appraisal of the studies included was not conducted, however this is not mandatory for scoping reviews.

## Introduction

The COVID-19 pandemic as initiated by SARS-CoV-2, was first reported in Wuhan, Hubei Province, in China in December 2019. Since then, globally, nations have been combatting it from a health and economic standpoint[1]. Due to its highly contagious nature, many individuals were forced into quarantine, with the potential for increased social isolation and loneliness among citizens, which has been linked to worse cardiovascular and mental health outcomes,[2]. Further, the SARS outbreak had a significant psychological impact on medical staff including the prevalence and rise of depression, acute stress disorder, alcohol dependency and post-quarantine mental distress,[3].

In the context of medical schools, as strict measures were put into place, medical schools which relied heavily on face-to-face teaching had to alter their traditional approach of teaching and learning activities towards online platforms,[4,5]. As a result, rapid changes were implemented, resulting in dramatic educational and potential psychological disturbances for medical students, particularly those in quarantine,[6,7]. This was further enhanced by the substantial academic coursework, the need to maintain their academic performance, and the potential for significant emotional stress among medical students. Further, coupled with the fear and uncertainty of the pandemic and its future course of action as depicted through a large media presence, this had the potential to take a greater toll on their mental and social wellbeing. A meta-analysis study by Moutinho illustrated that 34.6% of medical students suffered from depressive symptoms whilst 37.2% experienced anxiety symptoms prior to the SARS-CoV-2 outbreak, highlighting the significance of addressing and combatting the further negative impact of the pandemic on the mental health of these students,[8].

However, the impact of social isolation, necessitated by the quarantine required by medical students as a result of the COVID-19 pandemic is uncertain. Although it is tempting to assume an overall negative impact, some medical students may have been able to quarantine with family and friends which may have attenuated the negative impact of social isolation during the quarantine. Although there had been several individual primary studies addressing this topic, prior to starting our study, we were unable to find a systematic synthesis of the literature providing a comprehensive overview of the impact of quarantine on the psychological wellbeing of medical students during the COVID-19 pandemic. As medical students are our health care practitioners, a negative psychological impact of quarantine during the COVID-19 pandemic, is likely to result in anxiety and depression. Thus, if not addressed this creates long-term negative consequence on their overall quality of life and a long-term impact which may negatively affect the future quality of care given to the greater community. A scoping review of the literature was therefore conducted to identify the nature and extent of the available research evidence in this area. For this study, quarantine was defined as the period in which a person is kept in isolation to prevent the spread of a contagious disease.

## Methods

This study did not receive nor require ethics approval, as it does not involve human & animal participants.

The PRISMA-ScR (Preferred Reporting Items for Systematic reviews and Meta-Analyses extension for Scoping Reviews),[9] guideline, was used to structure this study. A systematic scoping review was carried out to explore the extent of published data across literature to assess the impact of COVID-19 on the psychological wellbeing of medical students. The nature of this review was crucial as it allowed for a systematic analysis and summarisation of appropriate information across various publications through its methodological framework,[10,11]. Further, this methodology was also guided by the utilisation of the PICOS table (Table 1) and a PRISMA-P 2015 checklist (Figure 1), allowing for a structured and focused approach to this study, based on the outlined inclusion and exclusion criteria.

**Table 1.** PICOS, inclusion criteria and exclusion criteria applied to database search.

| PICOS | Inclusion Criteria | Exclusion Criteria |
| --- | --- | --- |
| **Population** | Undergraduate and postgraduate medical students. | • Medical staff (including residents)<br>• Non-medical students<br>• General population |
| **Intervention** | Interventions through online questionnaires and phone calls. |  |
| **Comparison** | Comparisons of the various quarantine measures which have an impact on the mental wellbeing of medical students. |  |
| **Outcome** | Psychological impact of quarantine on medical students. |  |
| **Study design** | • Articles in English or translated to English<br>• All study designs including; mixed methods research, meta-analyses, systematic reviews, randomized controlled trials, cohort studies, case-control studies, cross-sectional studies, and descriptive papers<br>• Year of Publication: 1 January 2019 – 21 August 2021<br>• Databases: PubMed, Embase, ERIC, Scopus, Cochrane Database of Systematic Reviews, Web of Science |  |

**Figure 1.**
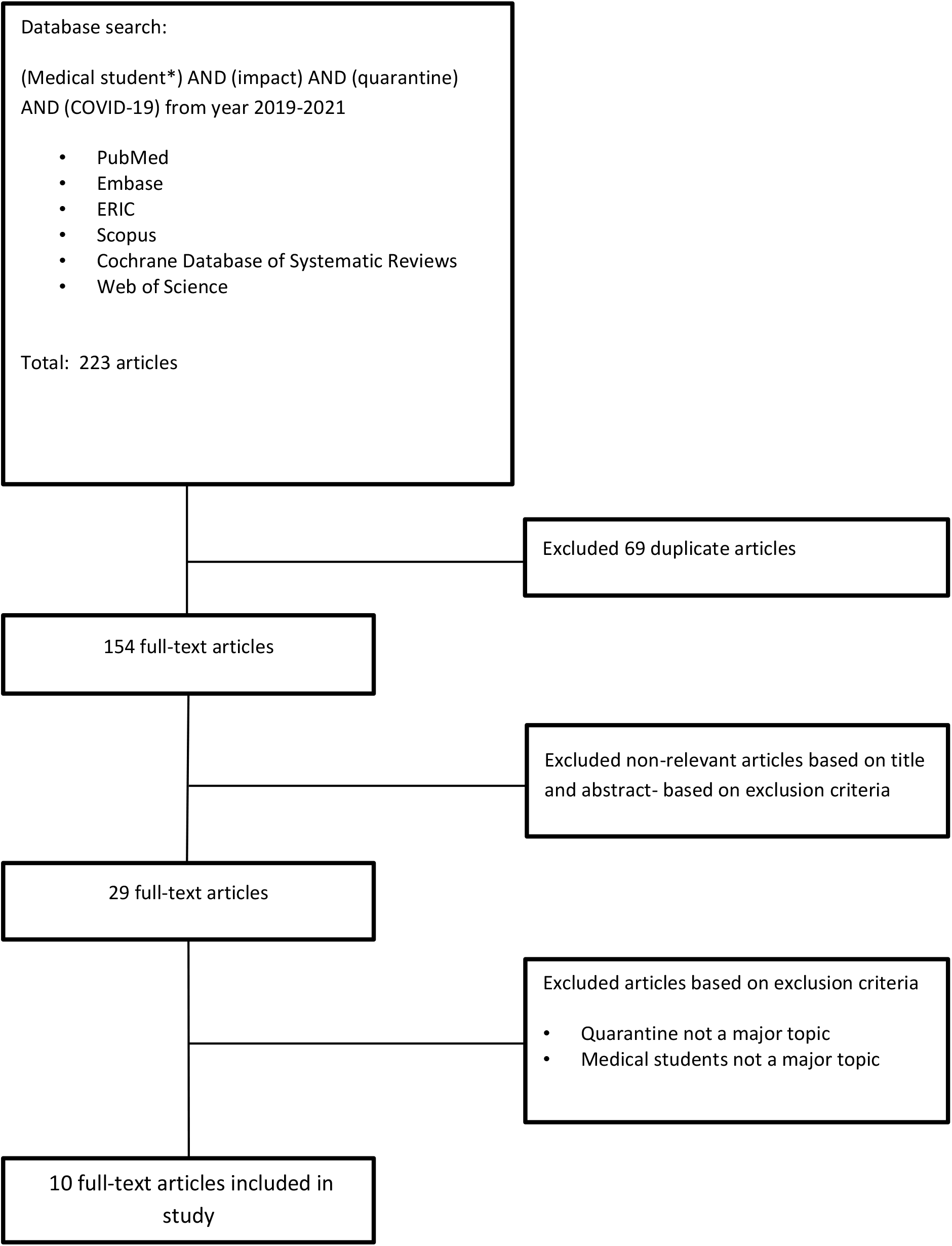
PRISMA flowchart.

To ensure a viable and appropriate research process, a search strategy was developed, consisting of the search terms, “medical student*” AND “impact” AND “quarantine” AND “COVID-19”. Searches were confined to articles published (excluding conference abstracts) between 1 January 2019-21 August 2021 on PubMed, Embase, ERIC, Scopus, Cochrane Database of Systematic Reviews and Web of Science as highlighted in Table 1. Further, for this study the population was limited to undergraduate and postgraduate medical students whereby data gathered was confined to either through online questionnaires and/or phone calls. Comparisons were evaluated between the various quarantine methods whilst analysing the psychological impact it had on medical students. Following this, a search of secondary references was conducted and despite finding articles addressing the impact of COVID-19 on medical students, no further articles were found which discussed specifically the impact of quarantine due to COVID-19 on medical students. All searches were carried out by the first author. Data from the selected studies was charted on a table on the Microsoft Word software and the following variables recorded; first author and year of publication, country of study, study design, sample size, focus group, mode of analysing impact of quarantine from COVID19 on mental health and results of study.

### Patients and public involvement

No patients or public were involved in the study.

### Results

A total of 223 articles were identified across the six databases, from which 69 duplicates were excluded resulting in 154 full-text articles. These articles were then reviewed based on their title and abstract in accordance with the exclusion criteria, resulting in 29 full-text articles. The full text of all 29 articles were then reviewed and screened again alongside the exclusion criteria, excluding those which did not include quarantine and medical students as major topics. Overall, 10 full-text articles were analysed in this study as displayed through the PRISMA process in Figure 1. Sample sizes ranged from 182 to 860 students and all studies used a variety of self-administered questionnaires to measure psychological wellbeing including depression, anxiety, stress, insomnia, and emotional stability.

Following a detailed analysis of the ten included articles, we found that the majority (8/10) of articles showed a negative impact of quarantine on the psychological wellbeing of medical students as seen through various self-administrated online tools and measurement scales (Table 2 and Table 3).

**Table 2.**
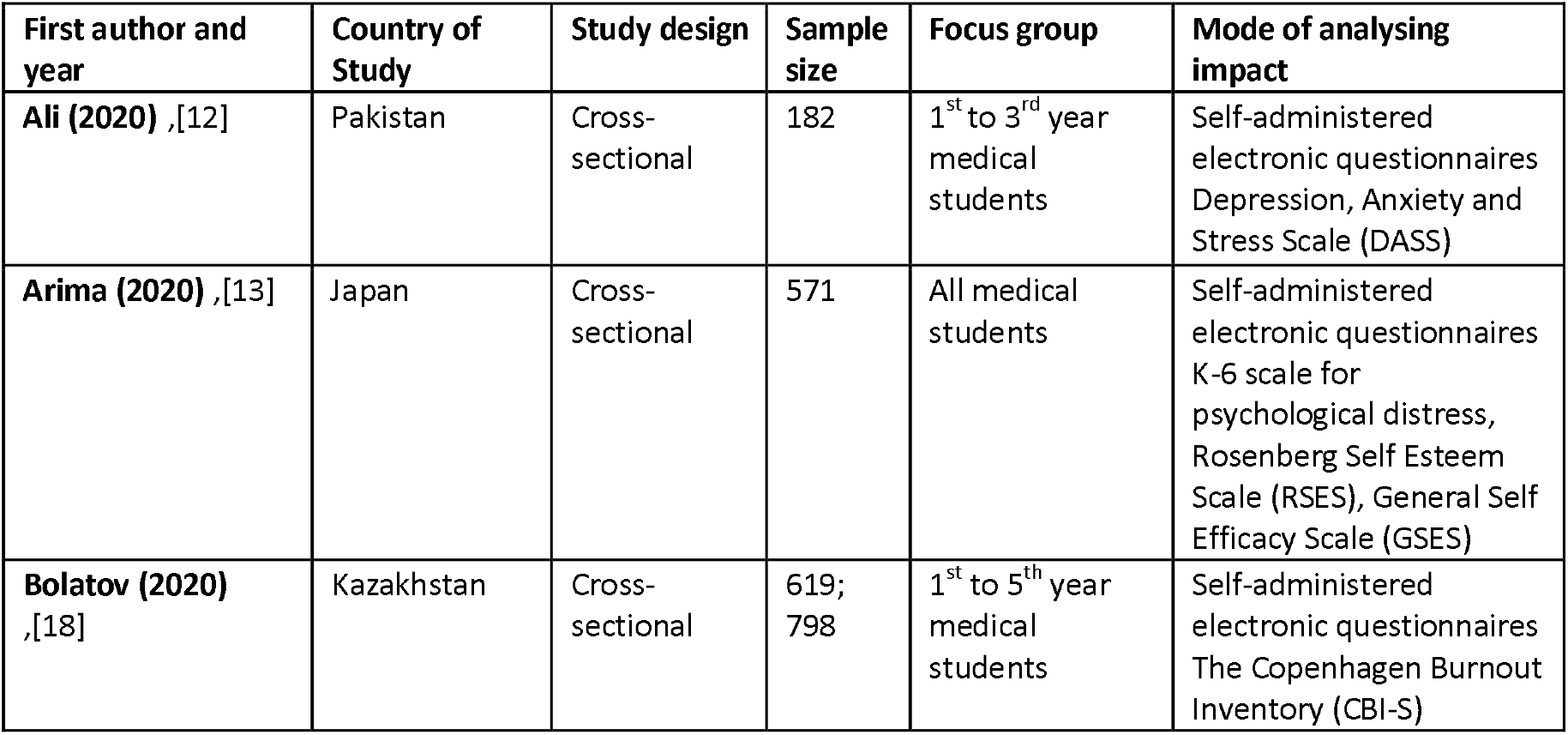
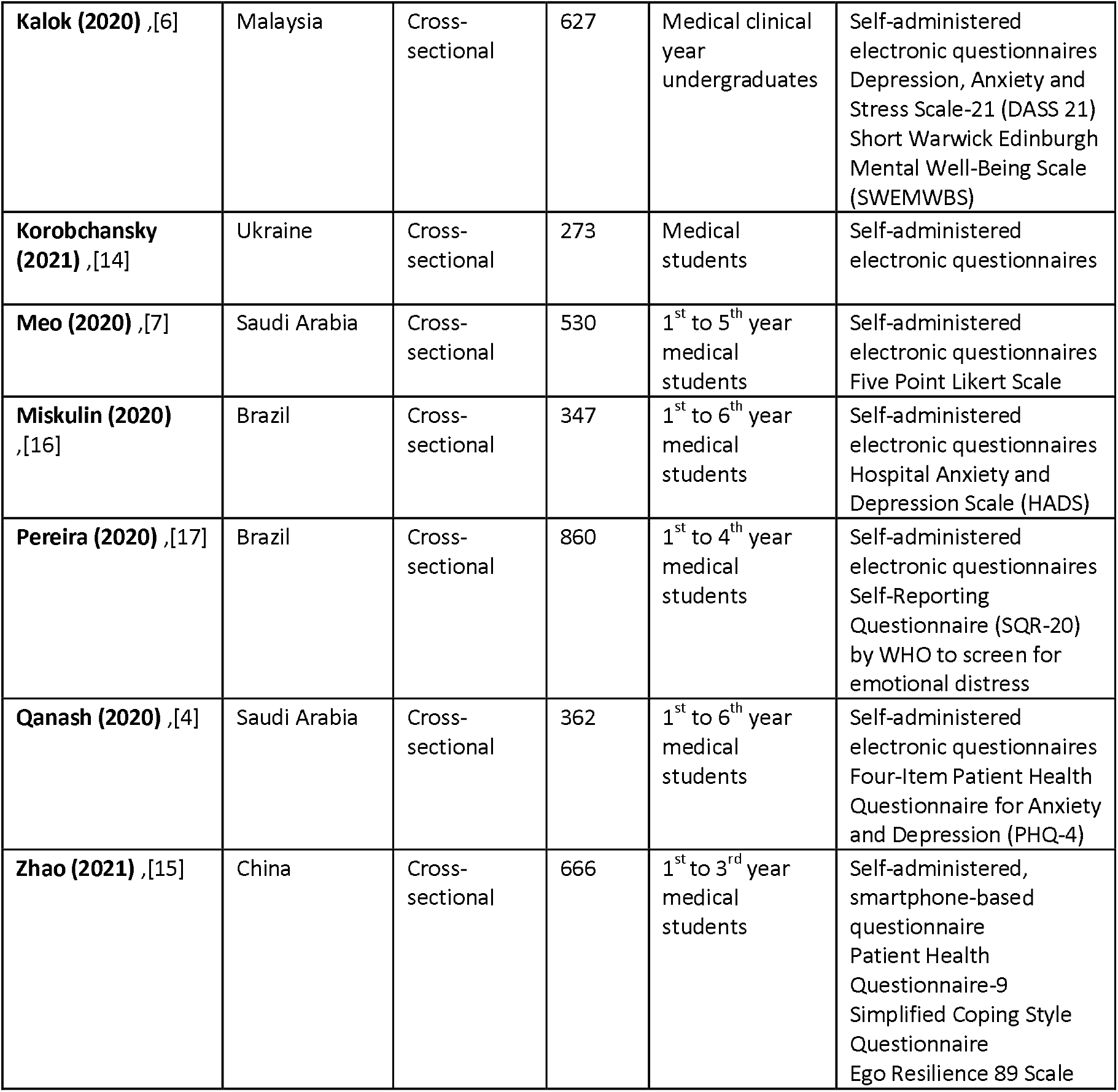
Summary of characteristics of included articles.

**Table 3.** Summary of findings of included studies.

| **First author and year** | **Results of study** |
| --- | --- |
| **Ali (2020)** ,[12] | Extremely severe depression- 38% of medical students<br>Depression levels were greater in first year medical students |
| **Arima (2020)** ,[13] | K-6 scale $\geq 5$ (significant degree of psychological distress)- 28.5% of medical students<br>Self-esteem & self-efficacy - influential factors in predicting psychological distress in medical students |
| **Bolatov (2020)**<br>,[18] | Depression during quarantine period- 27.6% of medical students; depression prior to quarantine- 49.3%<br>Anxiety during quarantine period- 15.5% of medical students; depression prior to quarantine- 42.3% |
| **Kalok (2020)**<br>,[6] | Negative impact on daily life, physical wellbeing & mental/ psychological wellbeing<br>Symptoms of psychological distress > 50% of medical students<br>Younger students (<25 years old)- more likely to experience anxiety<br>Increased social support- correlated with lower depression & stress<br>Family support & government support- independently associated with lower risk of depressive symptoms & greater mental wellbeing. |
| **Korobchansky (2021)** ,[14] | Deterioration of vision- 26.4% of medical students<br>Headaches- 33.7% of medical students<br>Absent-mindedness- 21.6% of medical students<br>Anxiety- 26.7% of medical students<br>Depression- 26.7% of medical students- depression<br>Sleep disturbance- 45.4% of medical students |
| **Meo (2020)** ,[7] | Depression- 23.5% of medical students<br>Experienced hopelessness, exhaustion or were emotionally unresponsiveness- 38.1% of medical students<br>Anxious or suffered from insomnia- 38.9% of medical students- |
| **Miskulin (2020)**<br>,[16] | HADS-D>8 (above cut-off)- 36% of medical students<br>First-year medical students had a greater prevalence (45.6%) of HADS-D>8 when compared to the other year medical students (32%) |
| **Pereira (2020)**<br>,[17] | No significant difference in common mental disorders (CMDs) between 2018 (62.2%), 2019 (60.9%) and 2020 (59.2%) for SQR-20≥7 (above cut-off) |
| **Qanash (2020)**<br>,[4] | Moderate to severe category in PHQ-4- 21.3% of medical students<br>Female and younger medical students were more negatively psychologically affected |
| **Zhao (2021)**<br>,[15] | Depression- 9.6% of medical students<br>11.1% of medical students who did not adapt to web-based classes had significantly higher depression scores |

The prevalence of depression among medical students was analysed across several studies; Ali depicting that 38% of students presented with extremely severe depression, Arima highlighting that 28.5% of students experienced a significant degree of psychological distress, whilst Korobchansky denoted a prevalence of depression in 26.7%; 23.5% through Meo’s analysis and evident in 9.6% of medical students through Zhao’s study,[7, 12 - 15]. Further, measurement tools analysing both anxiety and depression denoted that 36% of medical students had scores greater than the HADS-D>8 cut-off value and 21.3% were classified in the moderate to severe category in the PHQ-4 scale as highlighted by Miskulin and Qanash, respectively,[4,16]. Further, the presence of deterioration of vision, headaches, absent-mindedness, anxiety and sleep disturbance was studied alongside depression through Korobchansky’s analysis,[14]. Similarly, other factors such as presence of hopelessness, exhaustion, emotional instability, anxiousness, and insomnia were highlighted through Meo’s study,[7]. Arima explored the effect of self-esteem and self-efficacy as influential factors in predicting psychological distress among medical students,[7]. Other factors that were to be beneficial in combatting the negative impact of quarantine including the availability and extent of social support either in the form of familial or governmental support,[6]. As such, these factors were associated with a lower prevalence of depression and stress as well as a greater psychological wellbeing [6].

Contrastingly, Pereira’s longitudinal study illustrated a lack of significant variations in the prevalence of common mental disorders between 2018 (62.2%), 2019 (60.9%) and 2020 (59.2%) for the SQR-20≥7 cut off value,[17]. Further, Bolatov’s study unpredictably depicted that quarantine had a significant positive impact whereby both depression (27.6%) and anxiety (15.5%) rates decreased during quarantine when compared to the prevalence prior, depression (49.3%) and anxiety (42.3%),[18]. However, despite these two outlier studies, overall a substantial negative impact on the psychological wellbeing of medical students resulted due to quarantine during the COVID-19 pandemic, particularly amongst females and those in their early years of their medical degree,[4, 6, 12, 13, 16,].

## Discussion

The evidence is small but growing. Overall, the majority of studies analysed found a substantial negative psychological impact on the mental wellbeing of medical students as a result of the quarantine period during the COVID-19 pandemic. However, two of the ten articles reviewed showed varying results, with one showing no difference in psychological wellbeing and the other, a positive impact of quarantine on the mental health of medical students, which highlights the need for future comprehensive studies to further evaluate this research question.

A wide array of measurement tools and scales were utilized, as outlined in Table 2, highlighting the impact of a range of psychological symptoms including depression, anxiety, stress, insomnia and emotional stability as illustrated in Table 3. As such, these findings are significant as this study represents the first systematic scoping review that focusses on the impact of quarantine on medical students amidst the COVID-19 pandemic and as such sheds light into this field from which further avenues can be explored. Additionally, the negative psychological impacts that quarantine has had on these students are consequential, because, as future health care practitioners, anxiety and depression can result in long-term effects which impact the quality of care given to the greater community.

With respect to previous research, no previous articles were found and as such our understanding of the impact of quarantine on medical students is currently limited to the findings of this study. There is therefore a need for future analysis in this area. However, a study conducted across students at a Spanish University concluded that undergraduate students had significantly higher depression, anxiety and stress scores when compared to Masters students,[19]. Interestingly, this study also noted that Arts and Humanities students had the greatest anxiety and depressive scores when compared to their peers across other facilities,[19]. Further, another study conducted among French university students highlighted a high occurrence of severe self-reported depression, anxiety, stress, and distress as well as self-reported suicidal thoughts among those who were quarantined,[20].

The negative impact that quarantine has had on the psychological wellbeing of medical students could lead to long term health consequences as doctors, as depicted through studies that found an inversely proportional relationship between self-efficacy and depression related symptoms and how this can lead to the development of other negative psychological attributes,[21]. Thus, highlighting how this negative impact can lead to less resilience among medical students which consequentially results in challenging situations arising when faced with unfavourable circumstances. In contrast, Edwards’s study depicted that those medical students who possess high self-efficacy and self-esteem are more capable of handling various stresses, acquire better communication skills and interpersonal relationships which lead to an improvement in the physician-patient relationship,[22].

It is interesting to note that a study conducted in Australia during the equine influenza outbreak, depicted that individuals who were quarantined experienced greater levels of psychological distress compared to those who weren’t, emphasising the effect the associated isolation had as oppose to that caused by the outbreak alone,[23]. Thus, in our analysis, despite the two studies that strayed from the findings of the majority, our study has shown that quarantine during the COVID-19 pandemic, has had a significant detrimental impact on the psychological wellbeing of medical students globally. As such, it is vital to address these psychological issues to prevent further damaging consequences to both medical students and the community they serve. To address and tackle these negative psychological impacts, the implementation of wellbeing programs among medical students, effective contact tracing and mental health assessments should be considered. Further, another aspect is to focus on developing strategies to ensure medical students remain engaged, for instance through the implementation of interactive case discussions, and an increased engagement with pastoral support staff via online webinars.

### Strengths and limitations

The strengths of this study are associated with this study being the first systematic scoping review which encompasses findings from a range of universities globally. As such, this allowed for a comparison across institutions regarding their various methods of innovative online education teaching platforms as well as their associated negative impacts it has had on medical students. This is vital as it allows for a greater understanding of the various consequences that affect medical students. This is beneficial and significant for future studies which can aim to focus on developing methods to reduce future negative impacts on the mental health of students and ensure that the negative impacts that were caused are effectively managed and eliminated.

There were however, some limitations of this scoping review. Firstly, many of the studies analysed in this review were cross-sectional in nature and thus this potentially could have led to casual associations. Future studies should therefore, focus on being conducted in a longitudinal series. Secondly, because online questionnaires were used, different modes were used for analysing the data and student responses were mostly subjective. It was therefore difficult to accurately compare results across studies in a meta-analysis. Many of the studies also did not include a control group of medical students, pre-COVID-19 quarantine, highlighting the need for future studies to include two separate groups to allow for a contrast between the two, and a more accurate evaluation, albeit challenging to implement.

### Conclusion

Our scoping review found a small but growing literature on the impact of quarantine because of the COVID19 pandemic on the psychological wellbeing of medical students. All ten studies included the final review were cross sectional, used a variety of measurement tools and did not include any controls. Overall, the majority of studies (80%) found that the quarantine period during the COVID-19 pandemic had a negative psychological impact on the mental wellbeing of medical students. However, two of the ten articles reviewed showed varying results, with one showing no difference in psychological wellbeing and the other, a positive impact of quarantine on the mental health of medical students. This highlights the need for future comprehensive studies to further evaluate this research question. This is important to decrease and prevent the occurrence of psychological disorders among medical students. Therefore, in the meantime, it is recommended that medical schools implement targeted strategies and programs that aim to prevent and decrease psychological disorders among their students that may have arisen as a result of quarantine during the COVID-19 pandemic. By doing this, the potential long-term negative consequences on their overall quality of life may be reduced and the future quality of healthcare provided to the greater community, by these medical students, would be of safe and excellent standards as expected of medical students globally.

## Data Availability

All data produced in the present work are contained in the manuscript

## Author statement

Both authors met the four ICMJE criteria for authorship. WUA had the idea for the article, DV performed the literature search, DV and WA wrote the article, and WA is the guarantor. The corresponding author attests that all listed authors meet authorship criteria and that no others meeting the criteria have been omitted.

## Data Availability Statement

All data underlying the results are available as part of the article and no additional source data are required.

